# Lessons learned from a trial on integrated and proactive care for patients with complex needs (Hotspotters project): a process evaluation

**DOI:** 10.64898/2026.09.15.26362413

**Authors:** Vera Tiemes, Marije Verdonk, Mark van der Wel, Marieke Adriaanse, Laurens van Gestel, Marco Ephraim, Sytske van Bruggen, Marc Bruijnzeels, Mattijs Numans, Hanneke Borgdorff, Rimke Vos

## Abstract

**Background:** This study focused on adults with complex care needs due to problems on multiple life domains and consequent high acute care use. The Hotspotters Project was undertaken to assess the effects of proactive integrated care on health outcomes and healthcare costs in this group. We evaluated the reach, effectiveness, adoption, implementation and maintenance (RE-AIM) of the Hotspotters Project in order to identify factors that influenced implementation and outcomes.

**Method:** This mixed method study followed the RE-AIM-framework, using validated questionnaires, routine care data and semi-structured interviews. Qualitative data from patients and care providers offered insight in the quantitative outcomes. Quantitative data were analyzed descriptively (SPSS), and interview transcripts were analyzed using inductive thematic content analysis (Atlas.ti).

**Results:** As acute care use was lower than expected, few patients were eligible (153/1403) and subsequent low enrollment (n=30) limited conclusions regarding effectiveness. Nevertheless, the intervention improved collaboration with social work, and the structured intake was valued for the overview of patients’ complex needs. Many practices adopted the intervention into regular care, despite the complex inclusion process. Care providers proposed revising the target population and recruitment to better reflect the Dutch context.

**Conclusion:** Although cost-effectiveness could not be assessed, this study nonetheless provides insights that can help future interventions and trials to better match patients’ needs, adapt to local context and reduce patients’ study burden. Adapting the definition of ‘patients with complex needs’ to a local context can strengthen the rationale for implementation and enhance patient motivation.

## Introduction

Studies have reported that a disproportionately high share of care use may be attributable to a small group of patients(1, 2). One group of patients with high care use are the so-called ‘Hotspotters’. This population has complex care needs characterized by problems on multiple life domains and frequent use of acute care(3–5). These patients generally receive multiple forms of care, yet persist with relatively high care use over multiple years(2, 6), a large proportion of which is often potentially avoidable care(7). This suggests that current care for these patients may not fully meet their needs(3, 8). Moreover, these patients are less satisfied with the care they receive and show poorer health outcomes compared to other users of (acute) care(8–10). To increase treatment satisfaction, reduce preventable costs and improve health outcomes, it is important to address the unmet care needs of these patients.

In the Netherlands, as in many other countries, care for patients with complex needs is currently fragmented. This care also appears to be less effective –as indicated by high or potentially preventable care use –and can lead to frustration among all involved(3). Since healthcare and social care are organized and funded separately(11, 12), structural integration of social and medical care is limited. This is the primary explanation for the lack of overview and structured, easy access to resources or contacts with social care organizations often experienced by primary care providers(13–15).

The challenge of managing patients with complex needs, such as the ‘Hotspotters’ group, has long been recognized: between the 1960’s and 1980’s multidisciplinary patient-centered “home teams” or “health centers” were initiated to proactively assess and treat patients with ‘complex’ needs(16) –that is, medical needs that are affected by social vulnerability. These initiatives were well received on a small, local scale(17), but over time these teams were phased out due to a desire to centralize healthcare services(18).

Gawande’s 2011 *New Yorker* article on Hot Spotting, which described geographical clustering of high-cost patients(3), sparked renewed interest in case finding and integrated multidisciplinary or care/case management interventions(19). Initially, small scale studies showed promising outcomes(20), but later published reviews showed mixed results regarding care utilization, clinical and patient-reported outcomes(21–23). Mainly due to methodological study design issues (observational data with regression to the mean in a group with extreme care utilization), the evidence remained promising but not convincing(23, 24). In the absence of prior (cost)-effectiveness evidence we initiated the Hotspotters Project, a cost-effectiveness trial in Dutch primary care. This study offers proactive and integrated care to patients with complex care needs due to multiple life domain problems and high acute care use(25), employing a stepped wedge design to accommodate for regression to the mean effects(26, 27).

Since the Hotspotters project could be considered a complex intervention, a type of intervention that is challenging to implement(28), guidelines emphasize assessing how and under what circumstances a complex intervention brings about change(29) and the Hotspotters Project was evaluated as such in the present study. This study aimed to evaluate the reach, effectiveness, adoption, implementation and maintenance (RE-AIM)(30) of the Hotspotters project, both quantitatively and quantitatively, in order to identify factors that influenced implementation and outcomes. The lessons learned will be valuable for the future development of intervention programs (including cost-effectiveness) for patients with complex needs.

## Methods

This study reports the process evaluation of the Hotspotters Project for patients with complex care needs (protocol:(25)) following the RE-AIM framework(30). This study used a mixed methods approach, where qualitative data were used to provide insight in quantitative outcomes and lessons learned.

### Trial description

Complex interventions require some level of flexibility because strict standardization of the intervention can limit its implementation and effect(31). Therefore, in the Hotspotters Project we used a pragmatic approach concerning intervention components, provided these suited the intended purpose. The intervention started with an intake consultation visualizing issues on all life domains (Positive Health(32) or a similar tool(4D)(33)). Then, a personalized care plan was created in a multidisciplinary meeting with the physician, mental healthcare nurse, social worker and the patient (patient attendance was encouraged but not mandatory). A care coordinator provided proactive follow-up and supported care plan implementation (≥4 contacts). After obtaining written informed consent, all participants began with a control period followed by the intervention (12 months) and post-intervention follow-up. The length of control and follow-up phases depended on randomization (Figure 1). This study was approved by Leiden University Medical Centre ethics committee (METC-LDD, P21.123) and registered in clinicaltrials.gov (NCT05878054).

**Figure 1:**
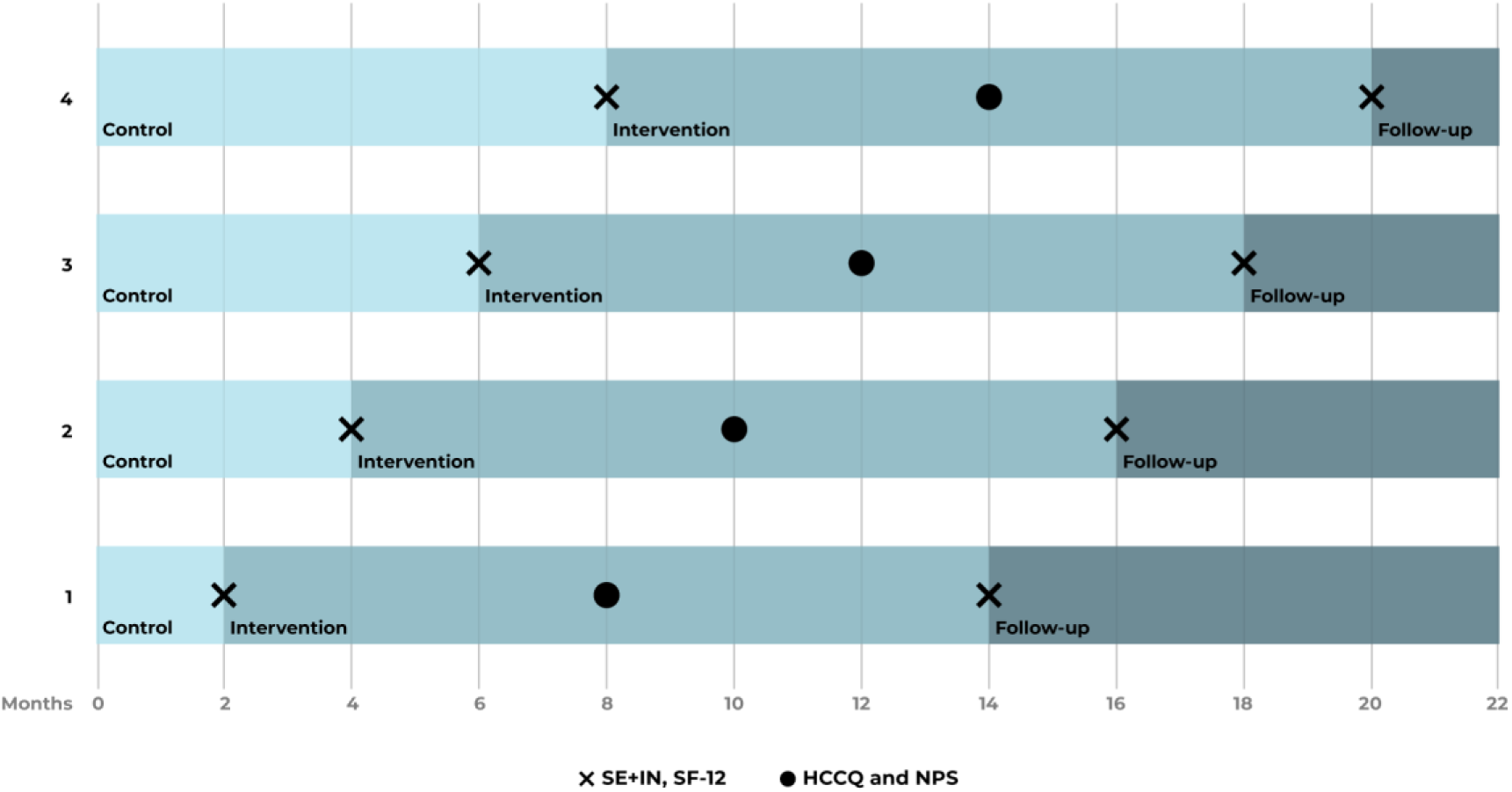
Trial-schedule: shows the duration of the control period (2-8 months), the intervention and follow-up (2-8 months). Primary care practices were randomized into one of four groups to determine the duration of the control period. The Self-efficacy and Intention item list (SE+IN) and the Short Form 12 (SF12) questionnaires were completed before the start of the intervention and 12 months later. The Health Care Climate Questionnaire (HCCQ) and the Net Promoter Score (NPS®) are administered 6 *months after start of the intervention. Not depicted: data collected for the cost-effectiveness assessment*.

Eligibility criteria were: problems in multiple life domains (physical, mental and/or social) and high acute care contacts (≥2, incl. out-of-hours primary care). Patients were sampled in two ways: by an Adjusted Clinical Groups-based algorithm(34) and by physician selection. Patients were then screened for eligibility and exclusion criteria. Eligibility assessments were conducted manually within each practice because automated screening was not feasible and privacy regulations applied. Practices contacted eligible patients to assess their interest in the trial, after which researchers provided study information and followed up to obtain informed consent.

### Implementation context and strategies

To understand why an intervention was implemented and how it functioned within the study setting, insight in the implementation context and provided strategies is needed, since it may influence the intervention delivery, uptake, and outcomes.

#### Implementation context

The Hotspotters Project was designed as a cost-effectiveness trial, which requires a control group, repeated questionnaires and clearly defined selection criteria. The definition of “high acute care use,” which varies in the literature from 4 to 24 emergency department visits per year(23), was set at ≥2 unplanned out-of-hour primary and/or acute care contacts in the previous year. This relatively low level of acute care use was chosen because lower rates were expected in the Dutch setting due to the gatekeeping role of primary care (mandatory before consulting a medical specialist) and the accessibility of comprehensive primary care, free of co-pay. Considering our target population had problems across multiple life domains, insight into their psychosocial wellbeing was considered important. However, this consequently increased the number of questionnaires and lengthened patient information leaflets. Because practice setup was labor-intensive, local eligibility screening— and thus trial initiation—was conducted sequentially per practice.

#### Implementation strategies

Implementation support was offered to practices during all trial phases, and the research team was available for additional support and to field questions from both the practices and patients.

#### Practice support

We offered staff support for eligibility screening. Practices received training in patient recruitment and were provided with a call script. All practices were offered (locally organized) training in Positive Health methodology. Near the end of the control period, practices received a reminder to plan intake appointments. When needed, we helped practices connect with local social work organizations. We also arranged additional funding to cover the extra time practices spent in multidisciplinary meetings. Practices received written and oral study instructions during a start-up visit, and prior to eligibility screening.

#### Target population support

We used several strategies described by Bonevski et al. to minimize barriers to study participation(26). Eligibility screening was carried out prior to recruitment, so all consenting patients could enroll in the trial. Practices were asked to consider oversampling. The inclusion period was extended to provide extra time to reach eligible patients. The stepped wedge design further guaranteed that all participants could receive the intervention, thus avoiding withholding a potentially beneficial program from a vulnerable patient group. With ethics committee approval, we tailored the study information to a low-literacy level and made it available in video format. Longer or more complex questionnaires were administered orally at first assessment. Phone support was made available for filling out questionnaires, both on request and when a questionnaire was not fully completed.

### Data collection

Administration logs were kept on the recruitment of care practices. Enrollment logs contained information on screening, recruitment and inclusion of patients.

#### Quantitative data

Care providers received a link to a questionnaire at the start of the trial and prior to the interviews. Patients received questionnaires digitally or, if preferred, by post. Routine care data and questionnaires were analyzed for all patients who completed the intervention and for whom data were available.

#### Qualitative data

Semi-structured interviews were conducted both with care providers and with patients. We invited all care providers in the trial for an interview, and all patients who completed the trial. Separate interview guides, consisting of open-ended questions, were created for primary care providers, social workers and patients (supplement B). Interviews took place face-to-face or via (video-)call. If preferred, care providers from the same practice were interviewed together.

### Outcome measures

#### Reach

The administration logs were used to assess the number of invited primary care practices that participated in the trial, while the number and proportion of eligible patients reached by the intervention was assessed using enrolment logs. The interviews provided insight into patients’ motivation for trial participation (Table 1).

**Table 1:** Dimensions of the RE-AIM framework applied to the Hotspotters intervention. *\*more details on effectiveness in supplement A*.

| RE-AIM dimension: | Evaluation among care professionals (data source) | Evaluation with participants (data source) |
| --- | --- | --- |
| **Reach** | Number of primary care practices invited to join the intervention compared to number of randomized practices (study administration logs). | Number of participants included in this study compared to the number of eligible patients (study administration logs/enrolment logs).<br>Motivation for participation (interviews). |
| **Effectiveness\*** | Perceived effectiveness (1-item questionnaire).<br>Outcomes of the intervention regarding patient health, appropriate care, and care use (interviews). | Change in primary care use (routine care data).<br>Experienced health (Short Form-12 questionnaire, SF-12).<br>Experienced care (Net Promoter Score).<br>Self-efficacy and intention (questionnaire).<br>Experienced health, appropriate care, and care use (interviews). |
| **Adoption** | Feasibility, appropriateness, and acceptability of the intervention (FIM, AIM, IAM questionnaires).<br>To what extent did it correspond with pre-existing vision or working methods? (interviews). | The extent to which it corresponded to pre-existing care (interviews). |
| **Implementation** | Fidelity of the care intervention. Role of care provider(s) and patients in multidisciplinary meeting. Extent of shared decision making (interviews). | Dose delivered: number of consultations for intervention (routine care data).<br>Experienced autonomy supportive care (HCCQ questionnaire).<br>Fidelity of the care intervention.<br>Patient's role in multidisciplinary meeting.<br>Extent of shared decision making (interviews). |
| **Maintenance** | Sustainment of (components of) the intervention after the trial, or application in other patients. Effect on collaboration between primary care and social care (interviews). | Sustainment of (components of) the intervention after the trial, application of insights/behavioral changes used in other situations (interviews). |

#### Effectiveness

Outcomes on effectiveness included questionnaires on self-efficacy and intention, health-related quality of life, care experience, and primary care utilization (Table 1, more details in *supplement A*).

#### Adoption

The anticipated and perceived feasibility, acceptability, and appropriateness were evaluated among the care providers using the following validated questionnaires: Feasibility Intervention Measure (FIM), Acceptability of Intervention Measure (AIM) and the Intervention Appropriateness Measure (IAM)(35). Each consist of 4-items (range: 1=completely disagree, 5=completely agree). Interviews explored whether care providers viewed the intervention as ‘new’ or as consistent with pre-existing care or vision (Table 1).

#### Implementation

The ‘dose delivered’ was calculated as the number of intervention consultations using routine care data. Patients’ perceived degree of autonomy supportive care was evaluated using the validated short form Health Care Climate Questionnaire (HCCQ, 6-items, range 1(worst) to 7(best)(36). Care providers and patients provided qualitative data on the use and form of the intervention components (Table 1).

#### Maintenance

Interviews with care providers explored whether they continued to use the intervention or its components, the patients targeted, and whether it had changed their collaboration with social workers. For patients, maintenance referred to sustained changes in health-related insights and behaviors following the intervention (Table 1).

### Analysis

Data from questionnaires (FIM, AIM, IAM and HCCQ) were analyzed using descriptive statistics (SPSS version 29.0). Missing or incomplete data were excluded (analysis regarding effectiveness in supplement A).

Audio-recorded interviews were transcribed verbatim using transcription software (Amberscript), reviewed by a researcher (MV/VT), and entered into Atlas.ti (v.24.0). Inductive thematic content analysis followed the steps of Braun&Clark(37), with organization of themes and codes guided by the RE-AIM domains. Analysis was performed by two researchers (MV, VT) and discrepancies discussed with a third (RV). Interview data were used to understand quantitative findings.

## Results

Despite strenuous efforts to support patient inclusion, the Hotspotters trial did not reach planned inclusion targets and was therefore terminated prematurely. Those already included in the trial were allowed to complete the (low-risk) intervention and full protocol. Five patients agreed to an interview, and care providers from 10 different practices were interviewed, consisting of 6 physicians, 4 mental health practice nurses, 3 social workers, 2 practice nurses, and 1 practice manager (*Supplement C)*. The interviews were conducted a mean of 13.7 months after start of the intervention and lasted on average 29 minutes. Experiences were generally consistent within practices but varied between practices. Differences by provider type are noted where relevant. The main results are summarized in table 2.

**Table 2:**
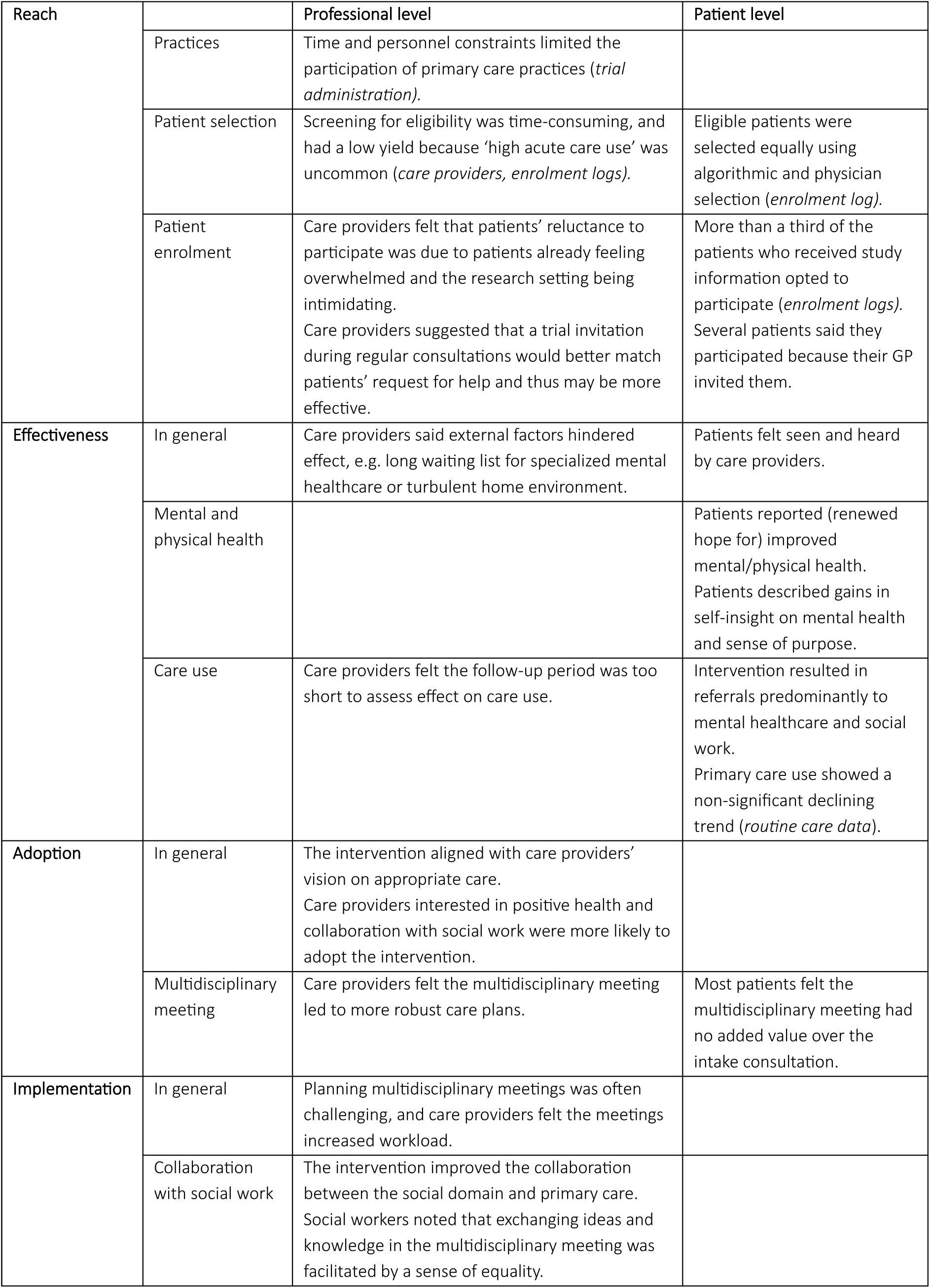
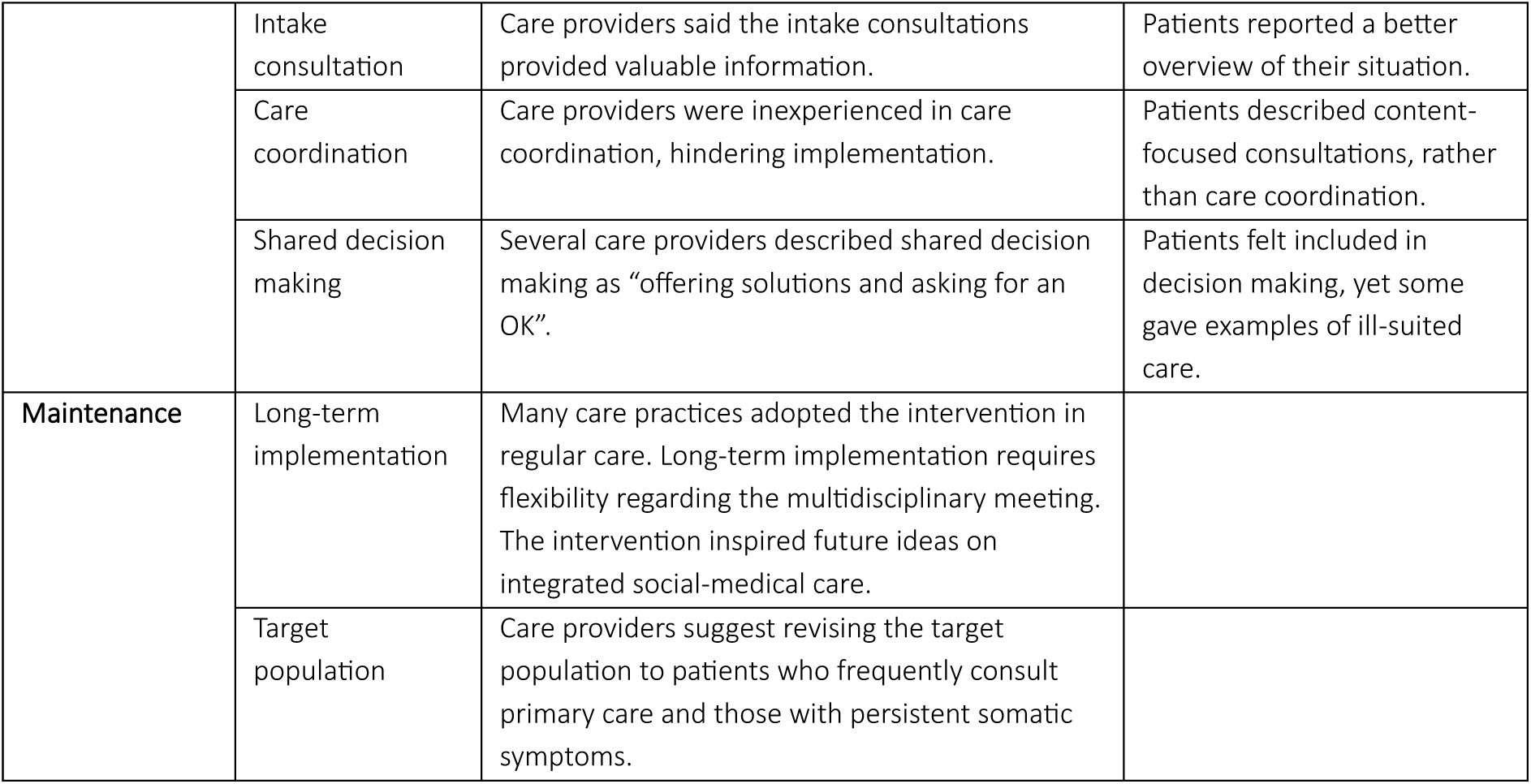
Overview of the main outcomes per domain of the RE-AIM. The outcomes derived from professional and patient data are presented separately. The majority of findings were derived from the interviews. If results were derived from other sources, these are noted in brackets.

### Reach

Fourteen out of 36 approached practices agreed to participate. The main reason for declining participation was time and personnel constraints. The intervention was not started in 5 of 14 practices due to no inclusions (n=2), withdrawal (n=2, personal constraints or issues with duration of control period), or due to premature termination of trial recruitment (n=1).

Reach of eligible patients was assessed in 11 of 14 practices, since screening information was unavailable in 3 practices (changed administrative requirements, linkage failure, or premature termination of trial recruitment). Eligibility screening and recruitment took more time and effort than care providers anticipated due to on-site patient selection and patients being contacted by the practice itself. Most patients (64.7%, n=908/1403) lacked high acute care use, resulting in extensive screening but a small eligible sample (n=153). Of those eligible, 81 patients consented to receive study information, of whom 30 gave informed consent (*see figure 2*).

**Figure 2:**
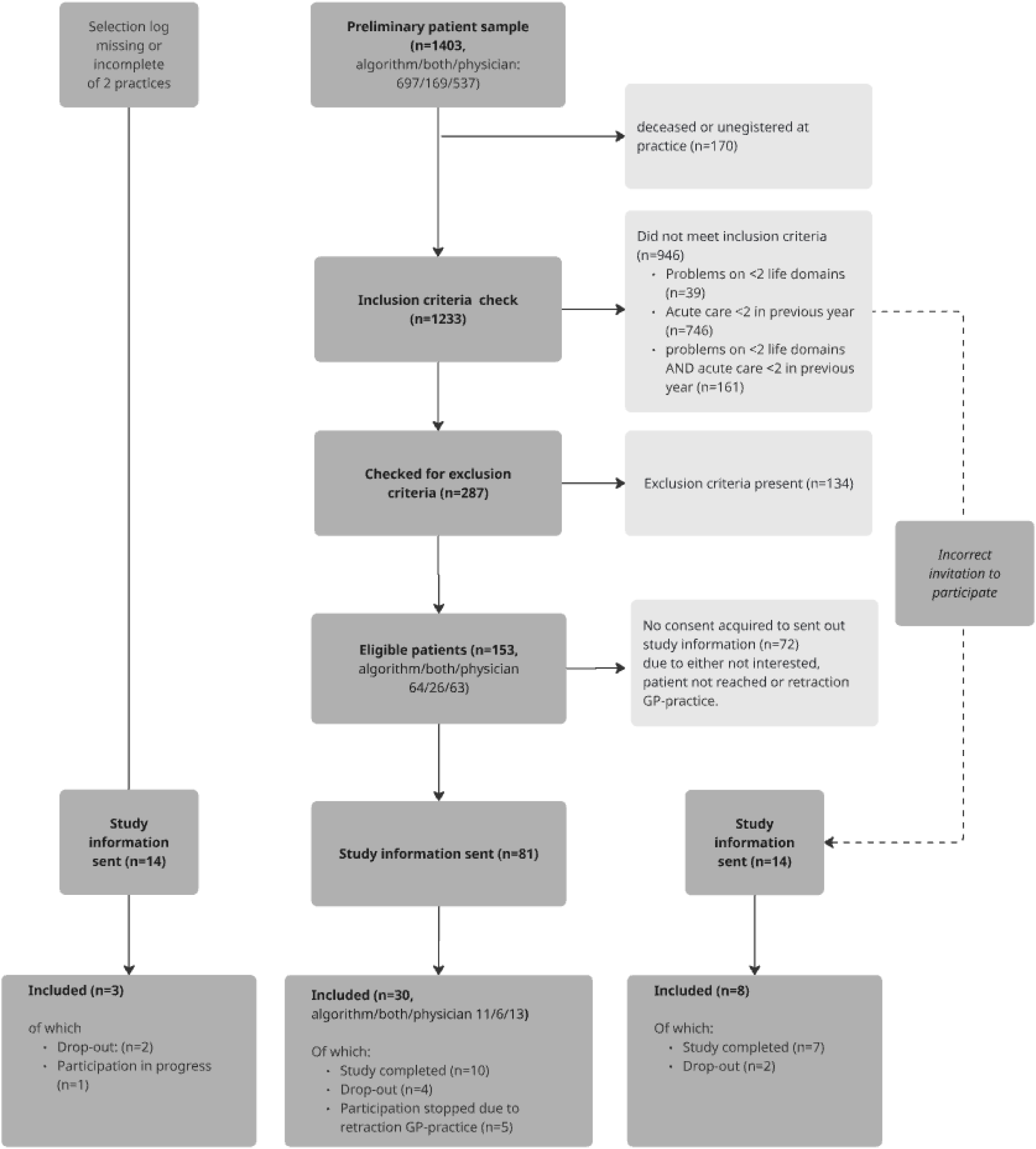
Flowchart of the Hotspotters Project screening and inclusion process. Reach was assessed in 11 practices with complete enrollment information (middle column). The preliminary sample consisted of 1403 patients screened for eligibility. The proportions of patients identified per method are reported as *algorithm/both/physician list. The left column concerns data from two practices with incomplete screening data, resulting in 3 inclusions. Incorrect invitation of (ineligible) patients is presented on the right. Not depicted in this flowchart: one practice where screening did not take place due to premature termination of enrolment*.

Care providers reported difficulty in reaching eligible patients and felt that many were reluctant to participate because they already felt overwhelmed in general or were intimidated by the research setting (prof-quote1). Care providers suggested that trial invitations during regular consultations would better match patients’ requests for help and thus might be more effective. Several patients participated because their GP invited them (pt-quote1). Some patients were under the impression that the study concerned evaluating and changing healthcare in general. This misunderstanding emerged in some practices during the intervention, while in others it was clarified early on and resulted in patients declining participation.

> Prof-quote1: “And, yes, actually there was a lot of ‘No, no, it’s not necessary. No thank you. No, it’s not needed.’ Various reasons: people who said ‘Oh yes, sounds interesting, but I really don’t have the time to do all that right now, and given everything that is going on privately…’
>
> Yes, so yes, basically there are people who did say yes, but then said ‘no’ again afterwards. […]”
>
> Pt-quote1: “We have two GPs and I don’t think much of one of them. I’m being very honest here, and the other one, well, she is a nice person. Yes, I can also tell her things now, […] and it was her who approached me and because it was her, I participated.”

### Effectiveness

Due to the low number of participants included, a full effectiveness evaluation could not be performed (more details in *supplement* A). Four of the five interviewed patients had positive experiences with the intervention, commenting that they felt seen, taken seriously, and had pleasant or helpful consultations. Patients indicated that the intervention provided personal insights on mental health and sense of purpose, and they experienced relief and support from discussing their issues, expressing a renewed hope of improving their situation. All interviewed patients reported that the intervention led to 2 or more referrals, predominantly mental healthcare (n=5), and to social work initiatives (n=4, e.g. sport, painting groups, volunteering). Care providers felt that intervention effects were hindered by external factors, like long waiting lists for specialized mental healthcare or patients’ turbulent home environment.

### Adoption

Care providers considered the Hotspotters’ intervention acceptable (median 4.0 pre- and post-intervention), feasible (median 3.25 pre- and 3.5 post-intervention), and appropriate (median 3.5 pre- and post-intervention) (n=23 and n=9, resp., supplement D). The intervention aligned well with primary care providers’ vision on providing appropriate care for non-medical problems. They were already interested in Positive Health, longer consultation times, and in collaborating with local care partners. Primary care providers perceived collaboration with social work, focus on patients’ strengths, and proactive care coordination as novel aspects of the intervention. One care provider noted similarities to solution-focused or integrated care approaches used in (specialized) mental healthcare (Prof-quote5, Prof-quote6).

Care providers commented that the multidisciplinary meeting contributed to more robust and jointly supported care plans. In one practice, however, the physician stated that discussing the patients’ goals identified in the intake felt ‘controlling’. Four of the interviewed patients were present at the multidisciplinary meeting, but only one of them felt this meeting had added value over the intake, improving understanding of symptoms and increasing motivation (Pt-quote6).

> Pt-quote6: “[on whether similar results would have been attained without a multidisciplinary meeting] Uh, yes, I… yes. It might not be a nice thing to say, but I think that it would have… gotten off the ground even then.”
>
> Prof-quote5: “[…] For me, positive health is actually almost synonymous with a previous methodology I had already worked with, the CRA method. With that method you also use a similar questionnaire, back then still on paper, to assess patient satisfaction across their life domains using scale questions from zero to 10.”
>
> Prof-quote6: “[on what was new] Well, of course, the setup was, wasn’t it? The combined start with GPs and a practice nurse together. So in that sense I think that the GP in particular was much better informed about the steps and about what I do with someone. That’s new, and of course, the involvement of the neighborhood social work team as well.”

### Implementation

The intervention improved collaboration with social work, although some care providers mentioned uncertainties regarding whether this was due to the Hotspotters intervention or to the simultaneously implemented initiation of social work in primary care. Social workers noted that exchanging ideas and knowledge in the multidisciplinary meetings was facilitated by a sense of equality. One practice substituted multidisciplinary meetings with social work referrals, due to difficulties in collaboration. The meetings presented some challenges regarding planning, with physicians mentioning increased workload and in two practices patients were not invited due to logistical concerns.

Patients (n=13) had a median of 5 intervention consultations (IQR 5, incl multidisciplinary meeting). All patients received an intake consultation. Care providers and patients valued the intake for providing an overview and for insightful information regarding the patients’ situation. All interviewed patients had recurring contact with their care coordinator after intake, but both patients and care providers described these contacts as problem-oriented rather than as care coordination (Prof-quote7). This was reiterated by one care provider, who noted inexperience and that maintaining a focus on care coordination required ongoing effort.

> Prof-quote7: “[…]GP5: “No, I agree, actually the same thing but with a slightly different approach in the conversation itself. But for the most part these were actually people I saw once every few weeks anyway, and then just mainly for stabilization. Yes, that is actually a bit of progress there… A continuation of that but taking a different approach in the conversations”

Care providers described the decision-making process as offering a solution to the identified problems and asking for an OK (prof-quote8). On autonomy supportive care (HCCQ) patients scored highly on the feeling that they were offered choices (median 6.0/7). Although interviewed patients said they felt included in decision making, some nevertheless gave examples of referrals to care that did not fit their needs, or for psychological help that required discussing topics that they felt uncomfortable with. One patient described that, while being asked for approval, an insufficient exploration of her concerns contributed to ill-suited care (pt-quote7).

> Pt-quote7: “Well, then I should have said no right away. Yes, and it was all properly asked and stated. I mean, yes, that’s it, that’s not actually the issue… absolutely not”.
>
> Prof-quote8: “Well yes, what you do is really ask everything about the domains and then see what’s there. And what kinds of interventions you could use based on that. And then you are specifically talking about social support or psychological support. And ultimately, yes, someone has to decide for themselves whether they want it. […] Yes, then you say, ‘It does seem like the things going on in your life are related to the fact that you often don’t feel well. Shall we do X or Y about that?’”

Support from the research team on practicalities, timing and study content was described as helpful, although some practices with a longer control period required additional support, as knowledge of the intervention steps had somewhat diminished during the interval between patient selection and intervention start.

### Maintenance

Several care providers expressed a wish for an on-site social worker to improve future integrated social-medical care. Of the 10 interviewed care practices, eight had adopted (part of) the intervention into regular care, including one with no trial inclusions. These adopting practices preserved the intake consultation, while adapting the multidisciplinary meeting and collaboration with social work to their context. Care providers revised the target population to frequent attenders of primary care, or patients where a non-physical cause of symptoms was suspected, thereby not considering acute care use (Prof-quote9, prof-quote10). Most providers considered the intervention less appropriate for patients with severe psychiatric conditions, as these patients often already received extensive care or lacked sufficient stability. Providers emphasized the importance of considering patients’ motivation and openness to non-medical approaches.

> Prof-quote9: “I really found it a difficult study for our population, …also due to the selection of the actual frequent attenders, the frequent users, not ours but from elsewhere. For various reasons. And it is quite difficult to explain to a person that we are going to do something for them because they often visit other centers. […] it is easier just to say “I see you a lot, don’t I? Let’s see if we can do something differently”. That seems a lot more appropriate than ‘you go to the emergency room or the GP out-of-hours service too often’.”
>
> Prof-quote10: “Persistent somatic symptoms is certainly a common denominator. But I wouldn’t do it only for PSS; I would also do it for people with mood disorders, that’s also really worthwhile. Loneliness, mood disorders, those aren’t always PSS sufferers …[].”

## Discussion

### Summary

The Hotspotters Project attempted to structure new opportunities regarding use of data and integrated social-medical care for patients with complex problems and high acute care use. We were not able to prove cost-effectiveness of the intervention, mainly due to low enrollment. Nevertheless, using a mixed-method process evaluation according to RE-AIM, we learned valuable lessons for daily practice and future research.

Few patients were eligible for inclusion –most lacked high acute care use –thus limiting conclusions regarding effectiveness. Care providers and patients valued the intake, which helped structure patients’ complex problems. The intervention improved collaboration between primary and social care. Care providers, unlike most patients, felt the multidisciplinary meeting contributed to more robust care plans matched to patient needs. It is, however, unclear how patients’ understanding of the meetings’ aims affected their experience. Care providers felt hindered by inexperience when taking on the care coordination role. Care providers proposed a revised target population and patient selection to better match Dutch ‘patients with complex needs‘, suggesting that this could increase patient motivation for participation and encourage care providers to adopt the intervention. Despite the trial’s complicated enrollment process, the majority of care practices continued to offer the Hotspotters intervention (or parts thereof) in regular care.

### Key findings and implications

Although intended to reduce costly acute care visits, the (cost-)effectiveness of The Hotspotters Project could not be demonstrated due to insufficient patient enrollment. This was mainly due to the finding that high acute care use is uncommon in the Dutch care setting, even with a lower threshold compared to international literature(7, 23). In our study, both screening methods predominantly identified patients with multimorbidity, rather than acute care use, suggesting that patients with complex needs may present differently across care systems. Care providers considered frequent primary care attenders and patients with medically unexplained symptoms as potentially representing ‘Hotspotters’ in the context of Dutch (primary) healthcare. Therefore, care providers recommended targeting these patients groups, which they felt better reflected patients’ help-seeking patterns and demand for primary care(38–40). Despite the relatively low cost of Dutch primary care, frequent attenders may nonetheless account for substantial healthcare expenditure through higher specialist care use and prescription costs(41). Future studies should assess the (cost-)effectiveness of the Hotspotters intervention in this population, as originally designed or as modified by adopting practices, and refine methods to identify patients at risk of persistent, frequent attendance. This would support practices target the intervention more effectively.

An interesting finding of the process evaluation was the adoption of the intervention into regular care by many practices, even in practices without formal trial inclusions. This was surprising, since evidence-based interventions are seldom implemented in practice beyond the trial period that includes implementation support(42): one typical example being the Collaborative Chronic care model for bipolar disorder that was not sustained in routine practice after implementation support ended, despite proven cost-effectiveness and nationwide implementation(43). The continued use of the Hotspotters intervention, despite difficult trial inclusions and the high trial-related workload, indicates that the intervention is perceived as appropriate and valued. Future implementations should retain the structured intake consultation, a component highly valued by patients and care providers, and some form of collaboration with social care. A notable finding concerned the multidisciplinary meeting for robust care planning, where there was a marked contrast between providers, who valued the approach, and patients, who generally felt that it had no added value over the intake consultation. Tailoring meeting timing and attendance to suit patient needs, while more actively engaging patients in meeting goals, may increase their perceived value. Furthermore, additional training may be needed to increase care providers’ confidence in care coordination.

The Hotspotters Project was designed to assess cost-effectiveness, to inform decision making regarding publicly-funded, large-scale implementation. Despite considerable effort and an extended inclusion period, enrollment remained low, underscoring the difficulty of conducting conventional effectiveness trials among patients with complex needs. Historically, researchers struggle to recruit and retain underrepresented populations, such as those with a socio-economic disadvantage, low (health) literacy or ethnic minorities(44). Despite using strategies to improve reach among hard-to-engage populations(26)—including a stepped-wedge design that ensures all participants receive the intervention, study information per video and plain-language formats, effect measuring in routine care databases, and support with questionnaire completion—care providers reported that the notion of participation in research was still perceived as burdensome. The complexity of these populations appears to limit the feasibility and validity of conventional RCTs. As a result, policy and guideline decisions for disadvantaged patient populations may need to be informed by a broader evidence base and a less restrictive evidentiary standard than is typically required. Future research may reduce the study burden by focusing on a limited set of clinically-relevant outcomes and leveraging routinely collected care data. In stepped-wedge trials, routinely collected care data serving as control data, can enable immediate intervention delivery upon enrollment.

## Strengths and limitations

The Hotspotters Project aimed to assess cost-effectiveness, with the ultimate goal of informed decision making regarding the implementation and reimbursement of these services. Despite the fact that cost-effectiveness could not be assessed, this process evaluation nonetheless provided valuable insights concerning the implementation and outcomes of the Hotspotters’ intervention that can support daily practice and help future research to develop new iterations of the intervention.

The evaluation included experiences from all parties involved –physicians, practice nurses, social workers, practice manager, and patients. This inclusion was essential, as illustrated by differing experiences regarding the multidisciplinary meeting and shared decision making. As these experiences were identified after data collection, it remains uncertain whether they could have been addressed through alternative approaches or clearer explanations.

Care providers from all practices were invited for evaluation, but not all participated. The few practices that were not interviewed may have had different experiences than those reported.

According to care providers, patients were reluctant to participate because they perceived the trial as an additional burden on top of their (already overwhelming) situation. It remains unclear whether this perception is indeed shared among non-participating patients. The pre-existing patient-provider relationship may have influenced how patients reported their reasons for non-participation. However, a more directly asking patients why they declined participation may be perceived as coercive.

## Conclusion

This process evaluation of the Hotspotters project found low inclusion rates that limited conclusions regarding (cost-)effectiveness. Unexpectedly, few patients met the inclusion criteria, mostly because high acute care use was less common than expected in the Dutch primary care setting. The main strengths of the interventions were an improved collaboration with social work, and a structured intake that provided an overview of patients’ complex needs. Surprisingly, despite low inclusion rates, many practices adopted the intervention into regular care. Furthermore, adapting the definition of ‘patients with complex needs’ to the local context is essential, and in the Dutch context a more explicit focus on frequent attenders in primary care would strengthen the rationale for implementation and enhance patient motivation by better matching needs and help-seeking patterns. However, this would still require cost-effectiveness analysis. Finally, the results of this process evaluation can guide research and practice in adapting to the local care context and help shape future iterations of the intervention to better match patients’ needs.

## Data Availability

The participants of this study did not give written consent for their data to be shared publicly.

## Acknowledgements

We thank the patients and care providers who shared their experiences with the Hotspotters Project. We appreciate the efforts of the steering committee of this project and all collegaes who contributed to this project. We thank Kimberley Leming for her contributions during the preparation and implementation phases of the Hotspotters Project. Her contributions included protocol development and Medical Ethics Committee procedures, recruitment and liaison with participating general practices, coordination of Positive Health training sessions for healthcare professionals, translation and administration of psychological questionnaires, data storage procedures, and preparation of the data for psychological outcome measures. Manual English editing was carried out by Dr. J-P. Bayley of MedicalEditing.nl.

## Supplement A: effectiveness

### Method

#### Outcome measures Effectiveness

Care providers scored expected intervention effectiveness pre-intervention, and perceived effectiveness post-intervention using a in-house-designed 1 item, 5-point scale (range: 1=completely disagree, 5=completely agree)(25).

An in-house-designed self-efficacy and intention item list was used to score patients’ intention to change and self-efficacy (when facing challenges) on four key behaviors (adequate self-care, maintaining daily structure, discussing concerns with care providers, asking for help in a timely manner). This 16-item questionnaire was administered pre-and post-intervention (range: 1=totally disagree, 5=totally agree)(25).

Effectiveness at the patient level was further assessed on health-related quality of life (HRQoL), experienced care and primary care use (*effect on care costs will be studied separately after all participants have completed the trial)*. Patients’ HRQoL was assessed pre- and post-intervention using the Short Form-12 (SF-12), a validated 12-item questionnaire(45). This was supplemented with qualitative data (*Do you feel healthier?).* Patients’ experienced care was assessed with the Net Promoter Score® (NPS®) on two items: the intervention and care coordination. The NPS® is a 10-point score that is commonly used to evaluate consumer or care experiences, and ranges from 1(worst) to 10(best)(46). This was supplemented with qualitative data on the care experience. Pseudonymized routine care data from primary care electronic health records were extracted using the Extramural LUMC Academic Network (ELAN)(47, 48). Changes in primary care use were evaluated by comparing average consultations per month at baseline, during the intervention and at follow-up. This was supplemented with qualitative data from care providers and patients on *new referrals* and *change in care use*.

#### Analysis

All questionnaire data, from both patient and care providers, were analyzed using descriptive statistics. One questionnaire, the Net Promoter Score (NPS®), is commonly calculated using a specific formula: percentage of promoters (those scoring 9-10) minus percentage of detractors (scoring 1-6). For easier interpretation, the NPS® results are presented both as the score according to this formula and as the median score (range 1-10). Missing or incomplete questionnaire data were excluded. For the SF-12, data were considered complete if pre- and post-data were present.

Change in primary care use was assessed by first calculating individual mean (non-intervention) primary care consultations per month at baseline (first 2 months of control period), during the intervention and at follow-up. These means were then used to assess the median number of consultations at baseline, intervention and follow-up (SPSS version29.0). The qualitative analysis is descripted in the manuscript.

## Results

Care providers’ perception of the effectiveness of the intervention remained unchanged (median 3/5 (IQR1)) (*table 4*). Care providers felt that intervention effects were hindered by external factors, like long waiting lists for specialized mental healthcare or a turbulent home environment (Prof-quote2).

Patients indicated that the intervention provided personal insights, for example regarding energy balance, uncovered trauma, and the need for hobbies or volunteering, as well as experiencing relief and support from discussing their issues, and renewed hope for improving their situation. One patient said the insights led them to pursue therapy and a change of work-life balance, resulting in better health and fewer primary care visits. Referrals sometimes led to unexpected effects. For example, one patient experienced negative outcomes after referral, feeling abandoned and denied help, whereas another patient found peer support for mental health through a referral to a sports club.

The Self-Efficacy and Intention item list showed no trend on motivation or self-efficacy (*table 5*).

> Prof-quote2: “[…] We also had a patient who was turned down several times in a row for a treatment that she actually … it was about regulation of emotion and she wasn’t accepted here, wasn’t accepted there… and every time she just fell short of meeting the conditions for the treatment setting and at a certain point this made her go: “okay, just forget it[..].”

### Mental and physical Health-related Quality of Life

The SF 12 showed a statistically non-significant increase in physical functioning and improvement in emotional role functioning (i.e., less interference with daily life) (*see table 5*). Two interviewed patients reported better physical and mental health due to receiving mental healthcare, a better overview of their situation, and overall improved experienced physical health. The other patients mentioned that their health remained unchanged, although one patient indicated finding renewed hope (Pt-quote2), and another noted that mental health issues may not resolve as quickly.

> Pt-quote2: “[on feeling healthier] No, I don’t think so. Just thinking. I do think I more strongly believe that there is still something that can be changed for the better. Yes, then I think, yes okay”.

#### Care utilization

Compared to baseline (2.0 consultations/month, IQR2.5, n=27), care utilization initially increased during the intervention (2.15 consultations/month, IQR1.8, n=27), but subsequently declined to 1.7 consultations per month (IQR 1.5, n=20) during follow-up. This change was not noticed by care providers. One patient mentioned visiting the GP less often, all others felt it remained unchanged –although some answered this along the lines of “still only when needed” (Pt-quote3). Two patients felt that care became more accessible.

During the intervention, all interviewed patients were referred to mental healthcare, consisting of a practice nurse (n=3), psychologist or psychiatrist (n=2). Other referrals concerned a lifestyle coach (n=1), elderly care specialist (n=1), physiotherapy (n=1), or social work (n=4, e.g. sport, painting groups, volunteering). Care providers considered these social work initiatives as supporting quality of life and thus appropriate care (Prof-quote3, Prof-quote-4).

> Pt-quote3: “No, no, [the GP visits] stayed the same. Only if there is really something wrong. Yes, and not just for any old reason. […]”
>
> Prof-quote3: “[one patient stated] “despite all my limitations, I find it so nice to think (despite all the problems) what else is actually possible?” So he really felt it was so helpful that someone else also wanted lend a hand, really get things going, he also started doing. Things were certainly set in motion, so I really think that is how positive health is meant to work. All of us have a positive feeling about it too.”
>
> Prof-quote-4: “[…] a man who’s had a lot of physical problems…. So we have…we really paid attention to his resilience, to his sense of meaning, to what he felt he needed and what made him happy, and that has also helped him with acceptance, so to speak. And the guy is now fully active; he is a volunteer with us. […] And he also started a living-room restaurant. We helped with the start-up and well, the guy has completely turned around, 180 degrees, which is very nice to see of course.”

#### Experience of care

Patients rated the care approach with a median 6/10 (IQR4), and care coordination 7/10 (IQR4). Most interviewed patients (n=4) had positive experiences with the intervention, stating that they felt seen, were taken seriously, and had pleasant or helpful consultations (Pt-quote5). One patient reported negative experiences with referred care due to a failure to keep promises and communication problems. This patient previously declined such referrals and described the referrals and subsequent negative experiences as ‘a risk of participating’. Another patient, although describing the consultations as pleasant, perceived these as non-contributory as they reiterated previous health insights.

> Pt-quote5: “[on consulting the practice nurse] Yes, that really encouraged me. Set me in a positive direction, so to speak. And that really helped a lot. […] it was a relief sometimes”

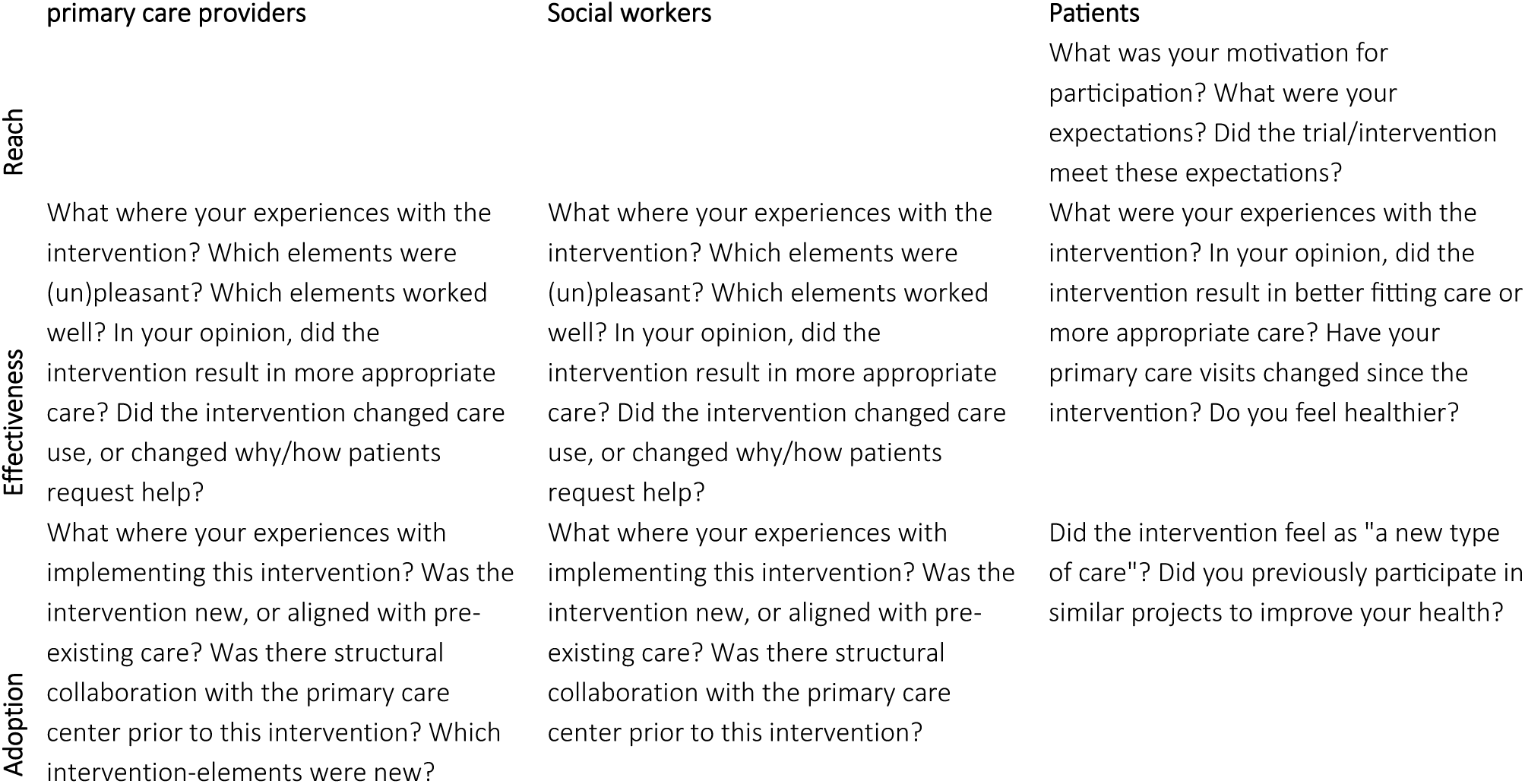
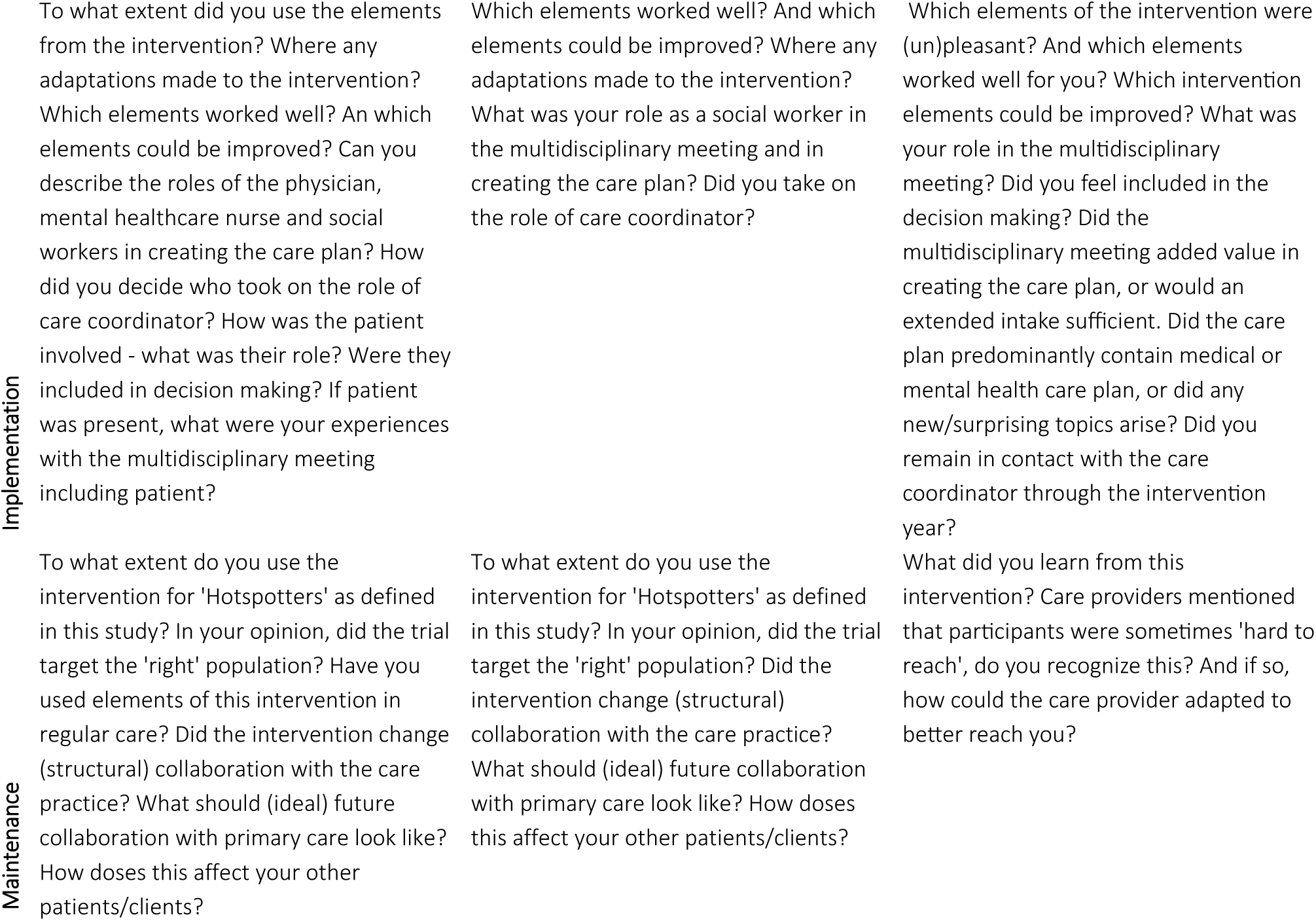

## Supplement B: interview guides

Supplement B shows the interview guides for data collection with primary care providers, social workers and patients. Both main questions and potential follow-up questions are included in this table.

## Supplement C overview of participants

**Table C1.** overview of interviewed patients.

| Patient | Sex | Age at inclusion<br>(ranges: 18-29, 30-39, 40-49, 50-59,, etc) | Practice |
| --- | --- | --- | --- |
| P1 | F | 70-79 | 1 |
| P2 | F | 50-59 | 2 |
| P3 | F | 70-79 | 3 |
| P4 | F | 60-69 | 3 |
| P5 | M | 60-69 | 3 |

**Table C2.** Overview of interviewed Care Providers.

| Care professional | Practice | Function | Number of included patients in practice (of which eligible) | Length of control period in months |
| --- | --- | --- | --- | --- |
| CP1 | A | GP | 6 | 4 |
| CP2 | A | Mental healthcare nurse | 6 | 4 |
| CP3 | B | GP | 2 | 4 |
| CP4 | B | Social worker | 2 | 4 |
| CP5 | C | Mental healthcare nurse | 10 (3) | 2 |
| CP6 | C | GP | 10 (3) | 2 |
| CP7 | C | Social worker | 10 (3) | 2 |
| CP8 | D | Mental healthcare nurse | 6 (5) | 2 |
| CP9 | D | Social worker | 6 (5) | 2 |
| CP10 | E | Practice manager | 0 | 8 |
| CP11 | F | Mental healthcare nurse | 3 | 4 |
| CP12 | F | GP | 3 | 4 |
| CP13 | G | GP | 2 | 4 |
| CP14 | H | Practice nurse elderly care | 4 | 2 |
| CP 15 | I | GP | 2 | 8 |
| CP 16 | J | Practice nurse | 1 | 6 |

**Table C3:** Sociodemographic data of patients in the trial.

|  | Patient invited for qualitative evaluation<br>(*trial complete*) | Patients included for quantitative evaluation<br>(*intervention complete*) |
| --- | --- | --- |
|  | N=11 | N=13 |
| Mean age in years (min-max) | 58,1 (32-78) | 58.9(32-78) |
| Female:Male | 7:6 | 6:5 |

## Supplement D: Quantitative results

**Table D1:**
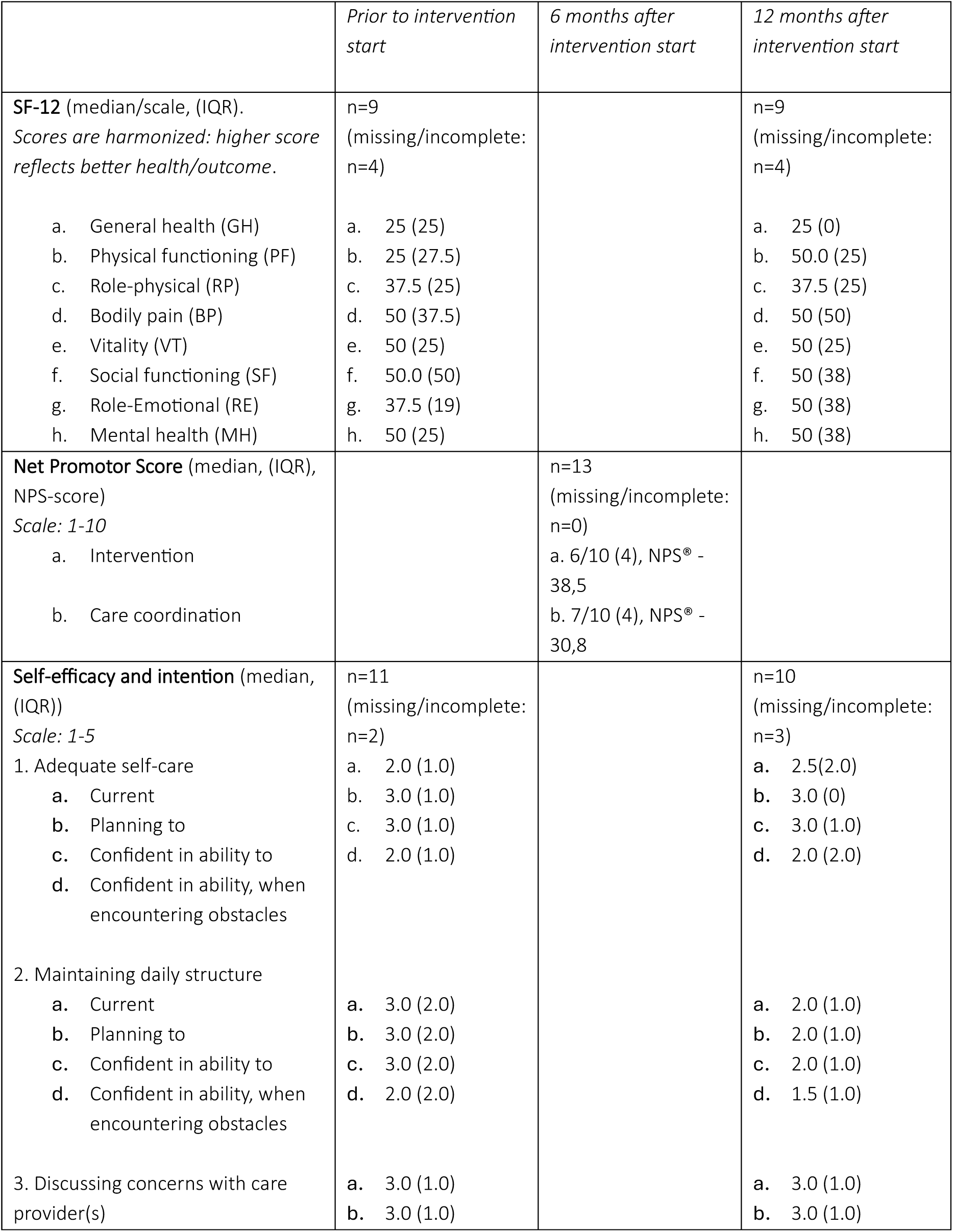
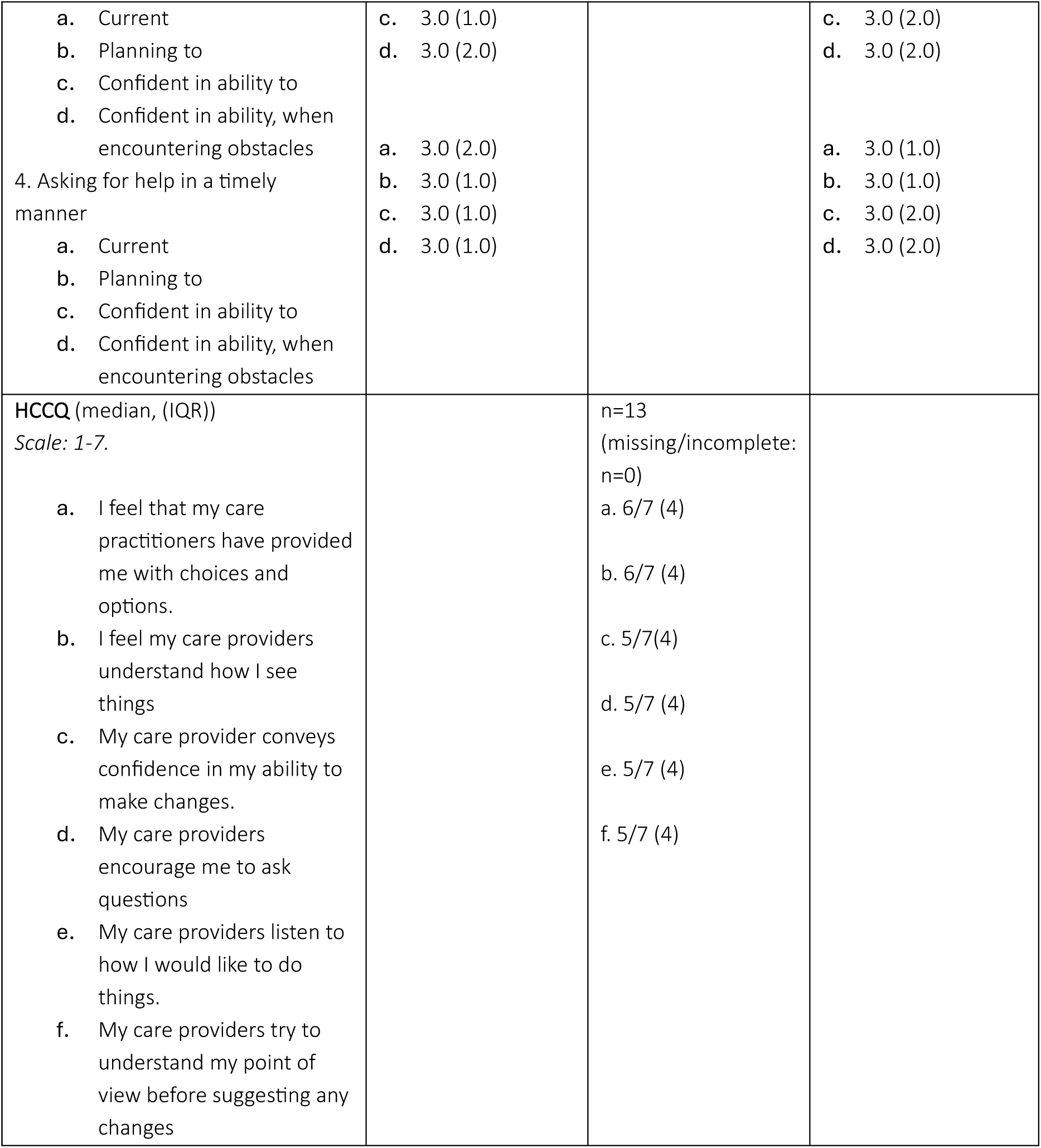
Outcomes of patient questionnaires on experienced health (SF-12), care experience (NPS), Self-efficacy and Intention, and autonomy supportive care (HCCQ). Per questionnaire, incomplete data was excluded. The results of the Short Form-12 (SF-12) were harmonized, meaning that for all items a higher score means better health outcomes.

|  | Prior to intervention start | 6 months after intervention start | 12 months after intervention start |
| --- | --- | --- | --- |
| **SF-12** (median/scale, (IQR)).<br>*Scores are harmonized: higher score reflects better health/outcome.* | n=9<br>(missing/incomplete: n=4) |  | n=9<br>(missing/incomplete: n=4) |
| a. General health (GH)<br>b. Physical functioning (PF)<br>c. Role-physical (RP)<br>d. Bodily pain (BP)<br>e. Vitality (VT)<br>f. Social functioning (SF)<br>g. Role-Emotional (RE)<br>h. Mental health (MH) | a. 25 (25)<br>b. 25 (27.5)<br>c. 37.5 (25)<br>d. 50 (37.5)<br>e. 50 (25)<br>f. 50.0 (50)<br>g. 37.5 (19)<br>h. 50 (25) |  | a. 25 (0)<br>b. 50.0 (25)<br>c. 37.5 (25)<br>d. 50 (50)<br>e. 50 (25)<br>f. 50 (38)<br>g. 50 (38)<br>h. 50 (38) |
| **Net Promotor Score** (median, (IQR), NPS-score)<br>*Scale: 1-10* |  | n=13<br>(missing/incomplete: n=0) |  |
| a. Intervention<br>b. Care coordination |  | a. 6/10 (4), NPS® - 38,5<br>b. 7/10 (4), NPS® - 30,8 |  |
| **Self-efficacy and intention** (median, (IQR))<br>*Scale: 1-5* | n=11<br>(missing/incomplete: n=2) |  | n=10<br>(missing/incomplete: n=3) |
| 1. Adequate self-care<br>a. Current<br>b. Planning to<br>c. Confident in ability to<br>d. Confident in ability, when encountering obstacles | a. 2.0 (1.0)<br>b. 3.0 (1.0)<br>c. 3.0 (1.0)<br>d. 2.0 (1.0) |  | a. 2.5 (2.0)<br>b. 3.0 (0)<br>c. 3.0 (1.0)<br>d. 2.0 (2.0) |
| 2. Maintaining daily structure<br>a. Current<br>b. Planning to<br>c. Confident in ability to<br>d. Confident in ability, when encountering obstacles | a. 3.0 (2.0)<br>b. 3.0 (2.0)<br>c. 3.0 (2.0)<br>d. 2.0 (2.0) |  | a. 2.0 (1.0)<br>b. 2.0 (1.0)<br>c. 2.0 (1.0)<br>d. 1.5 (1.0) |
| 3. Discussing concerns with care provider(s) | a. 3.0 (1.0)<br>b. 3.0 (1.0) |  | a. 3.0 (1.0)<br>b. 3.0 (1.0) |
| a. Current<br>b. Planning to<br>c. Confident in ability to<br>d. Confident in ability, when encountering obstacles<br>4. Asking for help in a timely manner<br>a. Current<br>b. Planning to<br>c. Confident in ability to<br>d. Confident in ability, when encountering obstacles | c. 3.0 (1.0)<br>d. 3.0 (2.0)<br>a. 3.0 (2.0)<br>b. 3.0 (1.0)<br>c. 3.0 (1.0)<br>d. 3.0 (1.0) |  | c. 3.0 (2.0)<br>d. 3.0 (2.0)<br>a. 3.0 (1.0)<br>b. 3.0 (1.0)<br>c. 3.0 (2.0)<br>d. 3.0 (2.0) |
| **HCCQ** (median, (IQR))<br>*Scale: 1-7.*<br>a. I feel that my care practitioners have provided me with choices and options.<br>b. I feel my care providers understand how I see things<br>c. My care provider conveys confidence in my ability to make changes.<br>d. My care providers encourage me to ask questions<br>e. My care providers listen to how I would like to do things.<br>f. My care providers try to understand my point of view before suggesting any changes |  | n=13<br>(missing/incomplete: n=0)<br>a. 6/7 (4)<br>b. 6/7 (4)<br>c. 5/7(4)<br>d. 5/7 (4)<br>e. 5/7 (4)<br>f. 5/7 (4) |  |

**Table D-2.**
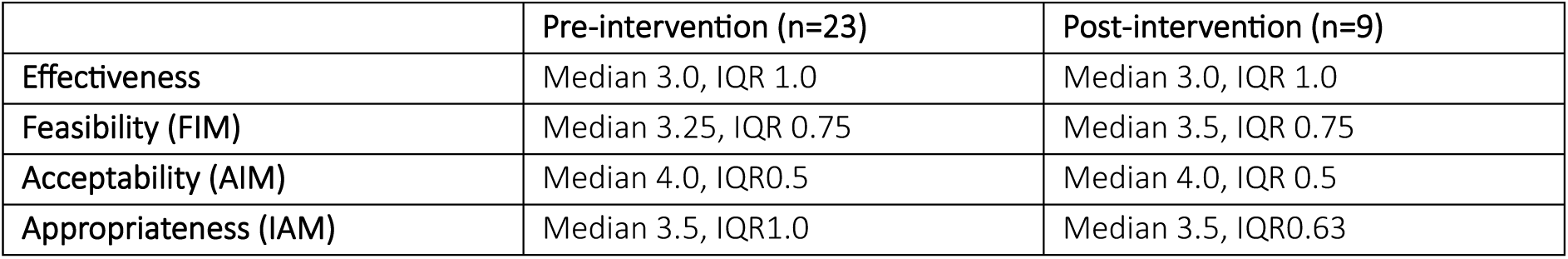
shows scores on Feasibility, Acceptability and Appropriateness as expected pre-intervention, and perceived post-intervention. The sample of pre-intervention and post-intervention partially overlap.

|  | Pre-intervention (n=23) | Post-intervention (n=9) |
| --- | --- | --- |
| **Effectiveness** | Median 3.0, IQR 1.0 | Median 3.0, IQR 1.0 |
| **Feasibility (FIM)** | Median 3.25, IQR 0.75 | Median 3.5, IQR 0.75 |
| **Acceptability (AIM)** | Median 4.0, IQR0.5 | Median 4.0, IQR 0.5 |
| **Appropriateness (IAM)** | Median 3.5, IQR1.0 | Median 3.5, IQR0.63 |

